# Unravelling Sex Differences in the Genetic Architecture of Anxiety

**DOI:** 10.1101/2025.02.09.25321968

**Authors:** Jihua Hu, Michelle K. Lupton, Enda M. Byrne, Nicholas G. Martin, David C. Whiteman, Catherine M. Olsen, Jodi T. Thomas, Sarah E. Medland, Katrina L. Grasby, Brittany L. Mitchell

## Abstract

Anxiety disorders show striking sex differences in prevalence, symptoms, and clinical characteristics, shaping how they manifest and are experienced. Here, we report the first sex-specific meta-analysis of genome-wide association studies (GWAS) of anxiety, leveraging two of the largest biobank datasets, UK Biobank and All of Us, comprising 85,014 female cases with 130,817 controls and 36,724 male cases with 117,044 controls. Functional annotation, sex-specific polygenic scores (PGS), and genetic correlations were performed to assess genetic differences and functional implications. In females, 26 lead SNPs were significantly associated with anxiety, compared to six in males. Among these, seven lead SNPs in females and five in males demonstrated significantly different effect sizes across sexes. Although the genetic correlation between sexes was high, it was significantly different from one, indicating partially distinct genetic architectures. In addition, both the SNP-based observed and liability-scale heritabilities (assuming a 2:1 female-to-male prevalence ratio) were significantly higher in females. Gene-based tests and functional prioritization identified different genes associated with anxiety in females and males. Moreover, genetic correlation analyses revealed stronger associations of female anxiety with attention-deficit/hyperactivity disorder (ADHD) and body mass index (BMI), whereas male anxiety showed stronger correlations with waist-hip-ratio adjusted BMI. While the overall genetic architecture of anxiety is largely shared, our findings reveal distinct sex-specific genetic associations and correlations, highlighting the value of analysing the sexes separately to uncover genetic signals that may be masked in sex-combined samples.

## Introduction

Anxiety disorders are the most common mental health disorders in the world, and a leading cause of disability-adjusted life years lost. Lifetime prevalence approaches one-third of the population, and demand for treatment is increasing, underscoring their substantial global public health burden (1).

Anxiety disorders are characterized by excessive and uncontrollable worry, persistent fear, and heightened perceptions of risk. Genetic factors have been estimated to contribute between 30% and 50% to anxiety disorders (2). Evidence from both twin studies (3) and genome-wide association studies (GWAS) indicates that much of this genetic liability is shared across anxiety disorders (4), including specific phobias, agoraphobia, social anxiety disorder, generalized anxiety disorder, and panic disorder (5). While the clinical diagnostic criteria vary for each disorder, the convergence of core symptoms and genetic liability supports examining anxiety disorders as a collective phenotype, referred to simply as “anxiety” in this study.

Striking sex differences have been observed in anxiety, particularly with regard to prevalence. Females are about twice as likely to be diagnosed as males (6). The prevalence difference between sexes has been consistently observed in specific anxiety disorders and across different cultures (7), and when all anxiety disorders are grouped together, the 2:1 prevalence ratio persists (8). Males and females also tend to manifest anxiety differently, with females commonly reporting more internalizing symptoms and comorbid depression, while males are more likely to exhibit externalizing symptoms and are more prone to substance use (7,9).

Previous studies have examined multiple underlying reasons for the observed differences in anxiety prevalence between sexes, with many focusing on gender-related influences such as masculinity norms that make men less likely to report anxiety (10), and more prone to using substances as a coping strategy (11). Studies focusing on biological sex differences have highlighted the potential roles of hormonal fluctuations (12), gut–brain axis interactions (13), sex-dimorphic brain activation during emotion processing (14,15), as well as genetic components. The contribution of genetic factors has primarily been examined through twin studies. Several studies have reported higher heritability in girls, including general anxiety symptoms (16), separation anxiety (17), and anxious and/or depressive symptoms (18). At the symptom level, genetic factor loadings on insomnia within the anxiety symptoms were stronger and more specific in females (19), and more recent work further demonstrated that anxiety itself was more heritable in females (20). While a large twin study of multiple anxiety disorders found that genetic contributions were highly similar in males and females (21). Overall, twin studies showed mixed findings, with some evidence for stronger genetic influences in females and sex differences in genetic effects.

To move beyond broad heritability estimates from twin studies, large-scale GWAS have begun to elucidate the molecular basis of anxiety, identifying many independent significant SNPs (ranging from 51 to 82 loci across studies) (4,22,23). However, none have focused on sex heterogeneity. In this study, we conducted the first sex-specific GWAS meta-analysis for anxiety to directly explore whether common genetic factors contribute to sex differences in anxiety. By examining sex-specific genetic associations with anxiety, we aimed to comprehensively characterize the genetic architecture of anxiety in females and males, and provide novel insights into the molecular basis of observed sex differences in these disorders.

## Materials and methods

### Study Population

This study consists of participants from two of the largest available biobanks, the UK Biobank (UKB) and All of Us (AoU) research program. UKB is a large nationwide cohort recruited across 22 centers in the United Kingdom from 2006, consisting of 488,377 genotyped individuals between the ages of 40 and 69 (24). AoU is an ongoing United States cohort recruited from over 340 sites, currently comprising more than 414,000 genotyped participants (25). Phenotypic information in UKB and AoU is derived from electronic health records (EHRs) and self-reported survey data (24,25). In both cohorts, we excluded individuals without genotype information, those with mismatched self-reported and genetic sex, and those who withdrew consent. This study included participants who were classified as white British within the UKB (26) and of European ancestry in AoU.

We identified participants who met DSM-5 criteria for the following anxiety disorders (5): agoraphobia, social phobias, specific phobias, generalized anxiety disorders, panic disorders and other phobic anxiety disorders as cases. In the UK Biobank, anxiety cases were defined by ICD-10, self-reported professional diagnoses, CIDI short-form criteria, or a GAD-7 score ≥ 8. Controls were participants without any self-reported or clinically recorded psychiatric diagnosis of mental distress. In All of Us, cases were defined by EHR diagnoses of generalized anxiety, panic, or phobic disorders, or by self-reported personal history of anxiety/panic disorder, while controls were participants with EHR and genetic data but no documented diagnoses of major psychiatric disorders. Case and control definitions in each cohort are detailed in the Supplementary Information **and** Supplementary Tables S1–S2.

### Genome-wide association analyses

Genome-wide association studies were performed using REGENIE (v 2.2.4) (27). Logistic regressions were conducted on chromosomes 1 to 23 in a combined sample, as well as in females and males separately. We assumed complete dosage compensation in females for testing associations of anxiety and variants in the non-pseudoautosomal region of chromosome X (genotypes in males were coded as 0/2). For UKB analyses, we adjusted for genotype array and the first four genetic principal components (PCs); for AoU analyses, we adjusted for the first 10 PCs. In the sex-combined GWAS, we additionally adjusted for genetic sex. Genetic variants with a minor allele frequency less than 1% or an imputation quality score less than 0.6 were excluded from the association test results.

### Meta-analysis

Meta-analysis of GWAS summary statistics from UKB and AoU was conducted using METAL ( v2020-05-05), using an inverse-variance weighted fixed-effects model. After meta-analysis, ambiguous SNPs and indels with inconsistent allele frequencies were excluded. The final meta-analysis comprised 369,599 participants, including 121,738 lifetime anxiety cases (85,014 females; 36,724 males) and 247,861 controls (130,817 females; 117,044 males) across both cohorts. LD Score Regression (LDSC) (v1.0.1) (28) was used to estimate the intercept as a check for inflation due to confounding from cryptic relatedness or population stratification.

### Characterisation of loci, functional annotation, and gene-based testing

To identify trait-associated variants within linkage disequilibrium (LD) blocks, we defined lead SNPs as those with r^2^ < 0.1 and P < 5×10^−8^ using FUMA(v1.6.1) (29).

Lead SNPs and their proxies (r² ≥ 0.6) were annotated in FUMA to prioritize genes, excluding the major histocompatibility complex region:

1. Positional mapping that mapped SNPs to genes within 10 kb;
2. eQTL mapping used data from the Brain eQTL Almanac (Braineac), GTEx V8 brain and adrenal gland datasets, focusing on significant eQTLs (FDR ≤ 0.05). The adrenal gland was included due to its role in the fight-or-flight response (30);
3. Chromatin interactions were mapped using all available Hi-C data in FUMA (FDR <= 1×10^−6^).

MAGMA (Multi-marker Analysis of GenoMic Annotation) in FUMA was used for gene-based, gene-set, and gene-expression association tests. Gene-based association, tested with the SNP-wise mean model, included SNPs within 2 kb upstream and 1 kb downstream of genes, with Bonferroni-corrected significance set at P < 2.493×10^−6^. Gene-set analysis explored shared biological functions using curated gene sets and gene ontology (GO) terms from MSigDB (v2023.1). Gene-expression analysis tested tissue-specific expression of associated genes using GTEx V8 data across 54 and 30 tissue types.

### Heritability and genetic correlations

SNP-based heritability was estimated using SBayesR (31), with a European-ancestry LD reference panel derived from the UK Biobank. SNP-based heritability was converted to heritability on liability (32) across a range of hypothesised population prevalences separately for females and males.

We investigated the genetic correlation between sex-specific anxiety and anxiety associated traits using LDSC, including mental health conditions, gastrointestinal symptoms, substance use, metabolism, some social behaviors, and sex hormones. Where available, we used both combined and sex-specific GWASs. All P-values were adjusted for FDR using the Benjamini-Hochberg approach. To determine significant differences between female- and male-specific genetic correlation with each trait, we calculated a Z-score as the difference between the sex-specific genetic correlations divided by the sum of the squares of their respective standard errors, and a two-tailed P-value from the Z-score (33); the corresponding adjusted P-value was computed using the BH method (34).

### Polygenic scores

To test whether our results were able to significantly predict anxiety in independent samples, we calculated polygenic scores (PGS) using SBayesR (31). SBayesR is a Bayesian technique that estimates the effect of SNPs from multi-normal distributions which may reflect the true distribution of genetic variants. The estimated PGS were standardized using the *scale()* function in R (v4.2.0).

We tested the association of PGS derived from our meta-analysis results with case-control lifetime anxiety status and current anxiety symptoms (GAD-7) in two population cohorts (QSkin (35) and PISA (36) respectively), as well as a large clinical, comorbid-depression cohort (Australian Genetics of Depression Study; AGDS (37)) (**Supplementary Information, Table 1**). While larger in size, it is important to note that in AGDS all the anxiety cases have comorbid depression. Therefore, this sensitivity analysis may distinguish prediction between population-based cohorts and clinical, comorbid-depression cohorts. To control for potential familial relationships, we conducted a restricted maximum likelihood (REML) analysis using GCTA (v1.91.7) (38), adjusting for the first 10 PCs and sex (combined analysis only).

**Table 1:** Sex Distribution in Cohorts Used for Polygenic Score (PGS) Predictions.

| **Cohort** | **Measurement** | **Nfemales**<br>**(cases/controls)** | **Nmales**<br>**(cases/controls)** | **Ncombined**<br>**(cases/controls)** |
| --- | --- | --- | --- | --- |
| Qskin | Self-report | 1,528/6,481 | 819/6,238 | 2,347/12,719 |
| PISA | GAD-7 | 3,263 | 1,605 | 4,868 |
| AGDS | Self-report | 8,278/6,481 | 2,902/6,238 | 11,180/12,719 |
| AGDS | GAD-7 | 5,556 | 1,681 | 7,282 |

To examine whether our PGS results were influenced by differences in sample sizes, and thus statistical power, across our discovery GWASs, we downsampled the number of cases and controls in the female-specific GWAS to match those in the male-specific GWAS for both UKB and AoU, thereby maintaining the male case–control ratio. These downsampled female GWASs were then meta-analyzed to obtain a downsampled meta-analysis result for females. In the prediction cohorts, we also downsampled female cases and controls to the same numbers as males. Downsampling was performed by random selection using the *sample()* function in R (v4.2.0). We then repeated the above PGS regression analyses using these downsampled results.

## Results

### GWAS of lifetime anxiety disorders in the total sample

We conducted a meta-analysis of 369,599 European-ancestry individuals for lifetime anxiety, including 121,738 cases. We identified 62 lead SNPs (**Table S3 and Figure 1A**) associated with anxiety. Among these, 35 were not in LD with variants reported in recent large anxiety GWASs (R2<0.8 or D’<0.9) (22,39). Using the same LD criteria and GWAS Catalog (40), seven were not proxies of known mental health–related variants. Quality metrics of each cohort GWAS are presented in Table S4, showing consistent SNP-based heritability with modest genomic inflation, and corresponding Manhattan plots for UKB and AoU are shown in **Supplementary Figures 1 and 2**. Cross-cohort LDSC analysis indicated high genetic correlation between UKB and AoU GWAS (rg=0.81 in females and rg=0.93 in males), supporting their combination in meta-analysis (**Table S4**). The sex-combined GWAS also showed strong genetic correlation with the previously largest anxiety study (rg=0.92, P < 1×10^-300^) (22). The observed heritability of lifetime anxiety was estimated at 0.12 (SE=0.002) (**Table S5**).

**Fig. 1:**
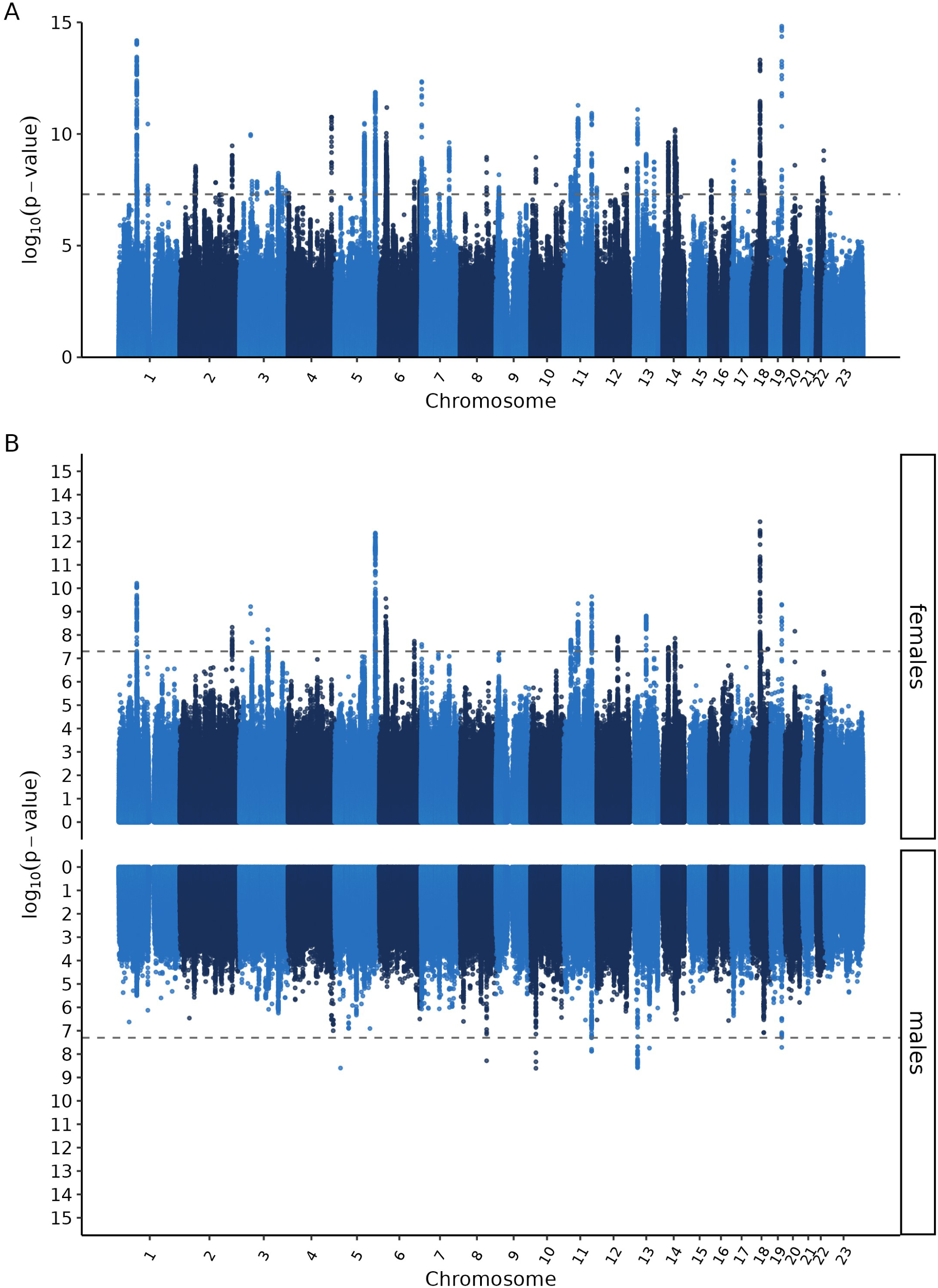
Manhattan plots of analyzed genetic variants for lifetime anxiety. A: Manhattan plot of anxiety disorders in the total sample (cases=121,738; controls=247,861); B: Miami plot of GWAS for females (cases=85,014; controls=130,817) are plotted above the X-axis, for males (cases=36,724; controls=117,044) are shown below the X-axis.

### Sex-specific GWASs of anxiety disorders

Sex-specific GWAS meta-analyses were performed using 215,831 (85,014 cases) females and 153,768 (36,724 cases) males. A total of 26 and six lead SNPs were identified in the female- and male-specific GWAS, respectively (**Table S6, S7** and **Figure 1B**). The strongest association in females was rs1000474011 (nearest gene: *CELF4*) on chromosome 18, and in males was rs113613364 (nearest gene: *NEBL*) on chromosome 10. A single locus on chromosome 19 showed evidence of overlap between females and males (R^2^=0.66), with no other shared lead SNPs identified.

Of the 32 sex-specific lead SNPs, 12 exhibited statistically significant differences in effect size, as determined by a Z-score test followed by FDR adjustment (**Figure 2**). Notably, one female lead SNP, rs73460081 (nearest gene: *CCDC102B*), showed an opposite effect direction between sexes, this SNP has also been prioritised to a redox-regulating gene *TMX3* based on chromatin interactions.

**Fig. 2:**
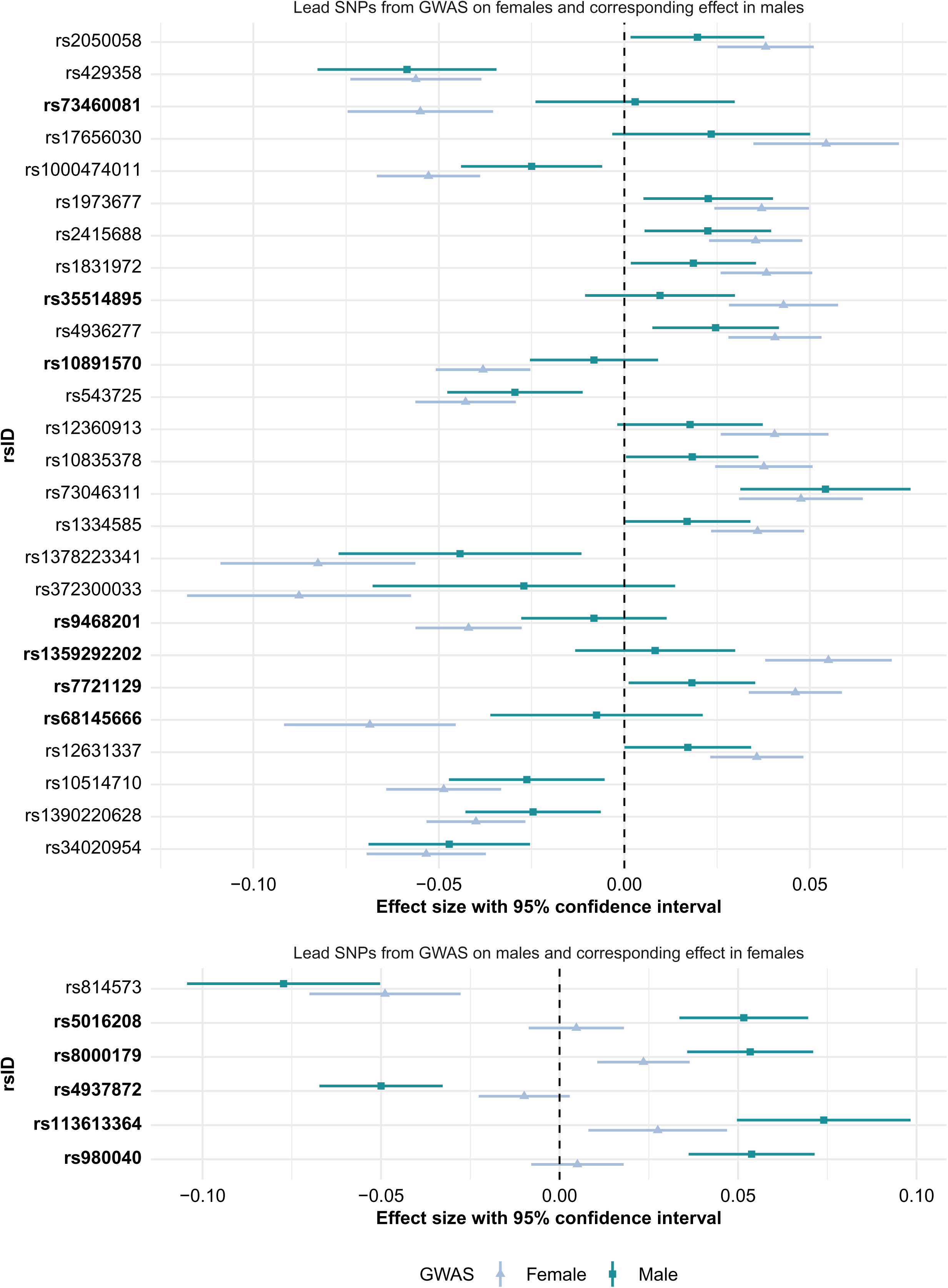
Forest plot of effect sizes and 95% confidence intervals of lead SNPs from the sex-specific GWAS. Effect size estimates (β) and 95% confidence intervals are shown for lead SNPs identified in sex-stratified GWAS. The top panel presents lead SNPs from the female GWAS with corresponding estimates in two sexes; the bottom panel presents lead SNPs from the male GWAS with corresponding estimates. SNP labels in bold indicate significant differences between female and male effect estimates, based on a Z-test for effect size differences with FDR-adjusted P < 0.05.

### Sex-specific heritability and cross-sex genetic correlation

SNP-based heritability of females was estimated as 0.14 (SD=0.003), and that of males was 0.12 (SD=0.004) on the observed scale. A Z-score test showed the SNP-based heritability was significantly different between females and males (Z=5.42, P=6.18×10^-8^). Males exhibited significantly higher genetic variance (Z=2.24, P=0.025), but also substantially greater residual variance, resulting in higher heritability estimates in females **Figure 3A (Table S5)**.

**Figure 3.**
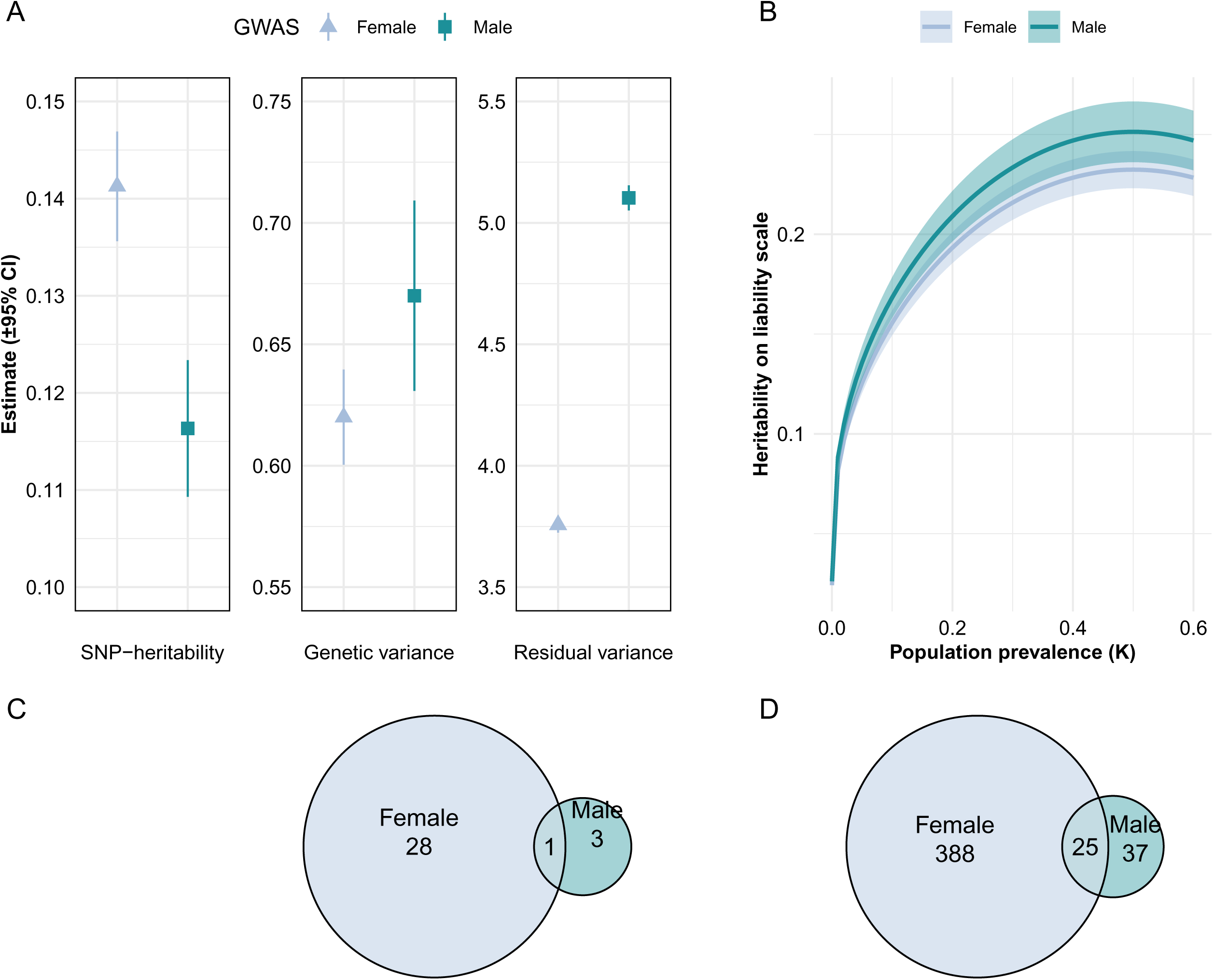
Sex-specific genetic architecture and gene overlap in anxiety. A: Observed SNP-based heritability and variance components (genetic and residual) estimated separately for females and males using SBayesR. B: Liability-scale heritability estimates across a range of assumed population prevalences (0–0.6) for each sex, based on observed-scale heritability and case proportions in each sex. C: Overlap of significant genes identified by MAGMA in females and males. D: Overlap of FUMA-prioritized genes across sexes.

To further evaluate sex-specific heritability in the context of varying population prevalences, we converted observed-scale estimates to the liability scale across a range of assumed population prevalences (0 to 0.6) (**Figure 3B**). When assuming the same population prevalence for both sexes, males showed significantly higher liability-scale heritability (Z=2.09, P=0.037), consistent with their greater estimated genetic variance (Figure 3A). In contrast, when assuming the population prevalence of females was 0.30 and of males was 0.15 as reported in the U.S (41), liability-scale heritability was higher in females (Z-test P=8.9×10^-4^).

The genetic correlation between females and males was 0.90 (SE=0.04), with the 95% confidence interval of 0.82 to 0.97 (**Table S5**). A t-test indicated the genetic correlation significantly differs from one (P=8.8×10^-3^), suggesting sex differences in the genetic architecture of anxiety.

### Gene-level analysis

To identify genes associated with lifetime anxiety, we used FUMA and prioritized 2,203 genes in the combined GWAS, 413 in females, and 62 in males (**Tables S8–S10**). Of these, 25 were shared between sexes (**Figure 3C**). Using MAGMA, 107, 29, and 4 significant genes were identified in the combined, female-specific, and male-specific GWAS, respectively (**Tables S11–S13**). Twenty-six female genes and three male genes overlapped with the combined results, with only one gene shared between the sexes (**Figure 3D**).

The metabolism-related genes *B3GALTL* and *APOC1* were both prioritized by FUMA and significant in MAGMA in males, and *B3GALTL* showed no association in females. A total of 24 genes were highlighted by both MAGMA and FUMA in the female-specific analyses. These included genes involved in neuronal signaling and synaptic function, immune-related butyrophilin family members, and several histone cluster genes. Gene-based scores from females were associated with 13 gene ontology (GO) biological processes of chromatin regulation-related gene sets, and olfactory transduction involved gene sets, while gene-based scores in males were associated with four GO terms related to lipoprotein clearance and efflux (**Table S14**).

### Polygenic score associations

Our combined anxiety disorder GWAS-derived PGS_C_ showed a strong association with self-reported lifetime anxiety in individuals in the QSkin population cohort (pseudo-R^2^ =4.6% and P = 7.54×10^-51^). When examining sex-specific PGS predictions, female-derived PGS_F_ showed the best prediction in both females and males (**Figure 4A, Table S15**). To exclude the potential influence of PGS derived from GWAS with differing power on the predictive ability, we downsized the GWAS for females and the combined sample to match that of males, and further equalized the number of females and males in the prediction cohorts.

**Figure 4:**
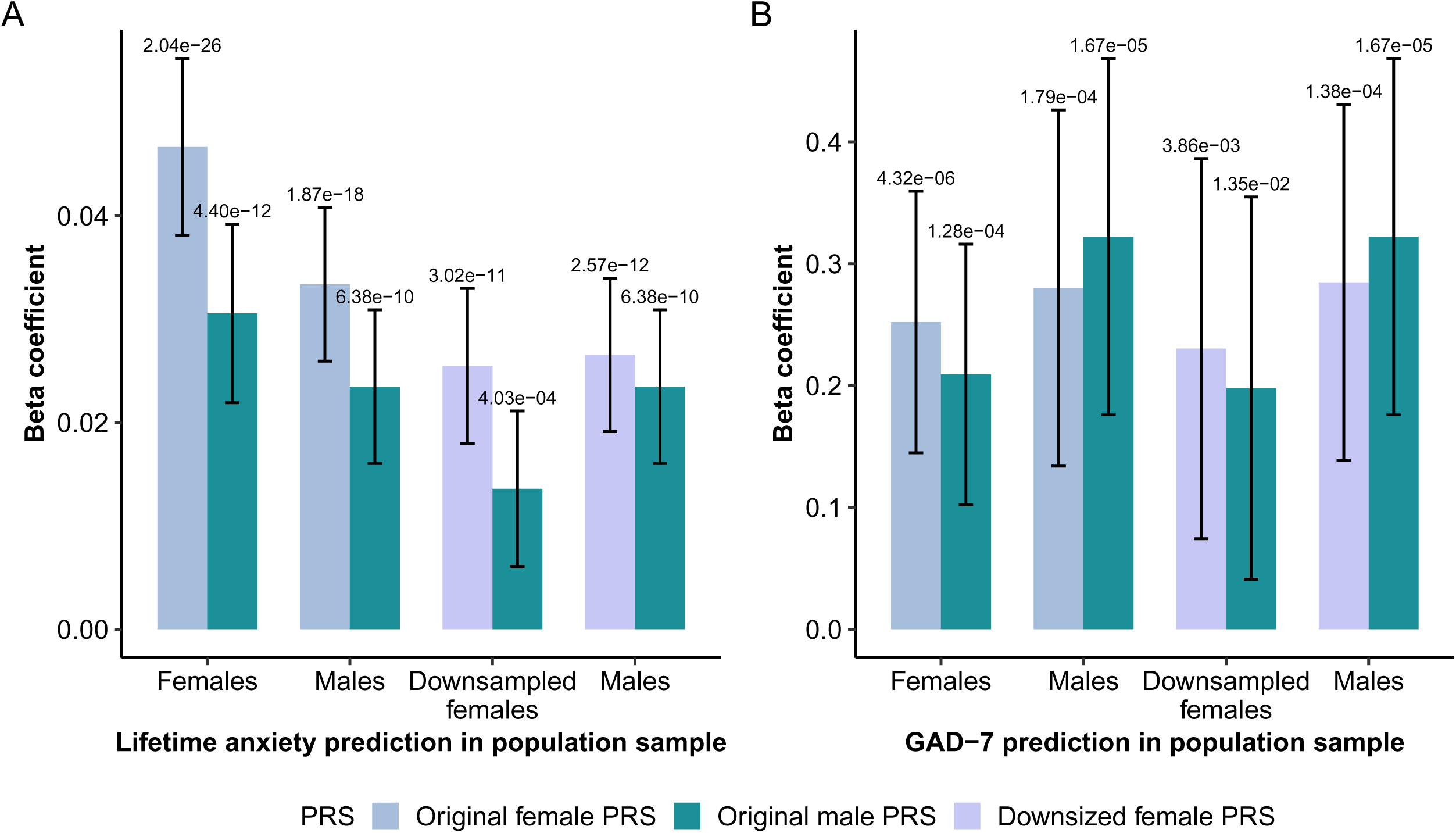
Sex-Specific Polygenic Scores (PGS) in Lifetime Anxiety and GAD-7. A: PGS prediction for lifetime anxiety in females and males in the population cohort (QSkin). B: PGS prediction for GAD-7 in the population cohort (PISA). Bars in different colors represent PGS derived from different GWAS. The X-axis indicates the sex of the target sample. P-values for the beta coefficient are shown at the top of the 95% confidence interval bar. To ensure comparability, an additional downsampled prediction was performed in which female-specific GWAS were downsized to match the male-specific GWAS, and the number of females in the target cohorts was also downsampled to equal that of males.

In the downsampled analyses of lifetime anxiety, the downsized PGS_F_ (pseudo-R^2^=1.6%) predicted significantly more variance in females than PGS_M_ (pseudo-R^2^ = 0.69%), as the Z-test comparing estimates yielded Z=2.19 (P=0.028). In males, the downsized PGS_F_ (pseudo-R^2^ = 1.7%) explained slightly more variance than PGS_M_ (R^2^ =1.4%), although this difference was not statistically significant.

We also evaluated predictions of current levels of anxiety using the GAD-7 score in another population cohort, PISA (**Figure 4B, Table S16**). Across both the original and downsized analyses, polygenic scores explained a comparable proportion of variance in males and females. In downsized females, PGS_F_ explained slightly more variance (R^2^*_F_* = 1.03%) than PGS*_M_* (R^2^*_M_* = 0.89%). In males, PGS_M_ predicted more variance (R^2^_M_ = 1.95%) compared to PGS_F_ (R^2^_F_ =1.70%). None of these differences in predictive value reached statistical significance.

Lastly, we tested the PGS associations in a clinical cohort where anxiety cases were comorbid with depression (**Table S17, Figure S1A**). The trend of the predictive ability of the original PGS was consistent with the QSkin and PISA cohorts. Once downsized, there was no difference in the performance of the PGS derived from the female-specific, or the male-specific GWAS when predicting into either sex (**Table S18, Figure S1B**).

### Sex-specific genetic correlations

We tested genetic correlations between our sex-specific GWAS and sex-combined GWAS across 28 traits (**Figure 5, Table S19**). Seven traits showed significant sex differences (Z-test P < 0.05). Females exhibited stronger correlations with attention deficit hyperactivity disorder (ADHD), metabolic syndrome, body mass index (BMI), and lower educational attainment, while males showed stronger correlations with bipolar disorder (all, type I, and type II). After FDR correction, the sex difference in ADHD remained significant.

**Fig. 5:**
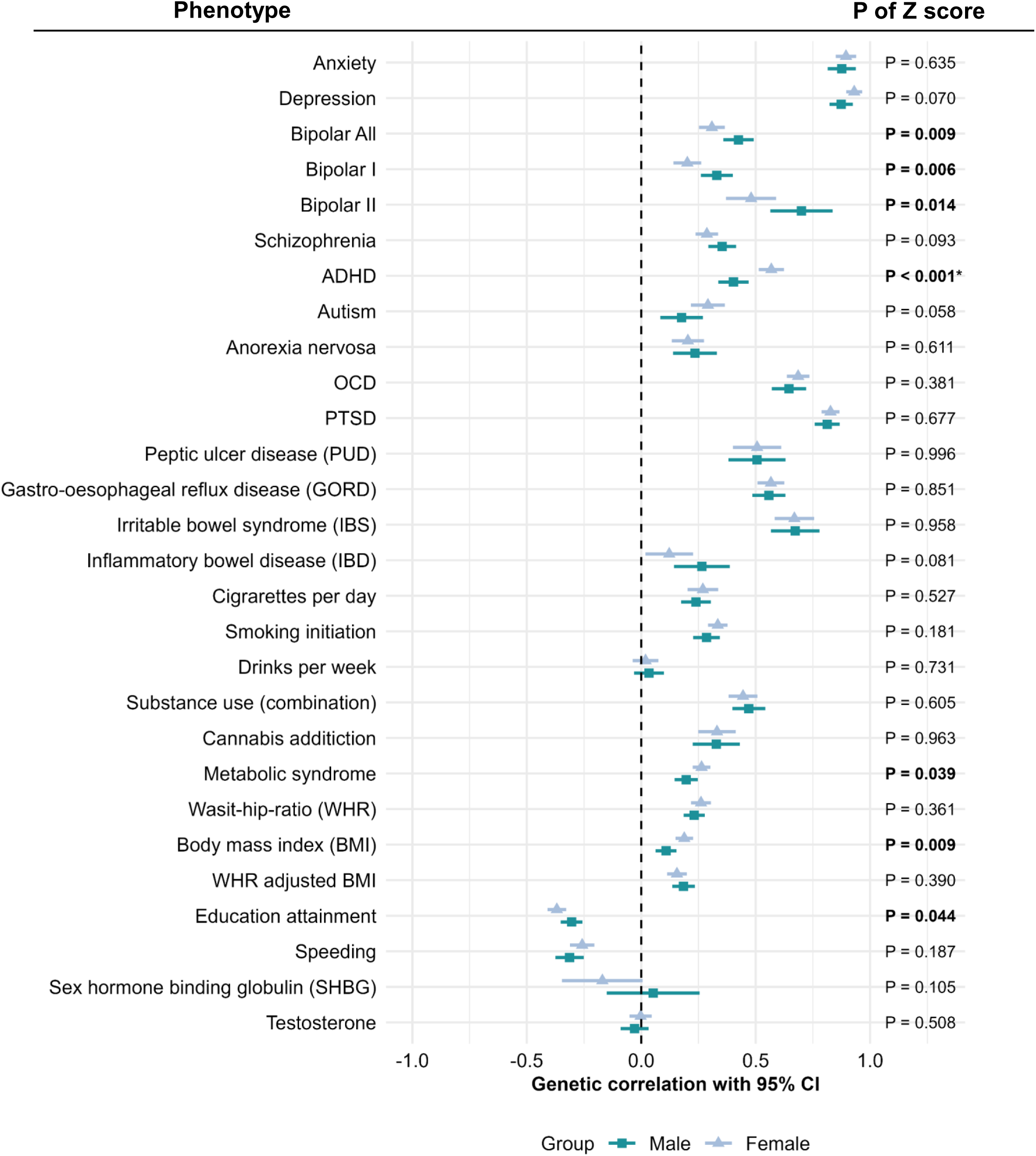
Forest plot of sex dependent genetic correlation. Genetic correlation (rg) estimates are represented by triangles (females) and squares (males). Horizontal bars indicate the 95% confidence intervals. P-values from the Z-tests comparing sex-specific rg estimates for each trait are shown on the right; significant values (P < 0.05) are shown in bold, and those passing the FDR threshold are additionally marked with an asterisk (*).

We further examined the genetic correlation of anxiety with available sex-specific GWAS of BMI and waist-to-hip ratio adjusted for BMI (WHRadjBMI) (**Table S20**). The stronger genetic correlation observed between females and BMI was maintained and remained significantly greater than that in males (Z=2.42, P=0.015). For WHRadjBMI, which did not differ by sex when using sex-combined GWAS, stratified analyses revealed a stronger correlation in males (Z=3.05, P=0.002).

## Discussion

Our study set out to investigate the genetic basis of sex differences in anxiety disorders by leveraging sex-stratified GWAS and including the often-overlooked X chromosome. To our knowledge, this is the first study to comprehensively examine potential sex-specific genetic influences on anxiety disorders using this methodology.

Through sex-specific GWAS, we observed differences in significantly associated SNPs and annotated genes between females and males. Seven female-specific and six male-specific lead SNPs showed statistically significant differences in effect size between the sexes. In addition to differences in specific SNPs, female- and male-stratified GWAS revealed very different prioritized genes and corresponding biological processes. Genes prioritized in females were enriched in chromatin regulation and olfactory transduction, while genes prioritized in males were involved in the regulation of lipoprotein levels. Chromatin regulation, including nucleosome remodeling, chromatin condensation, and histone methylation, influences gene expression and epigenetics (42). Histone modifications have been linked to anxiety, stress responses, and fear memory retrieval (42). The enrichment of chromatin interaction-related GO processes in females, but not males, may suggest greater environmental sensitivity. Olfactory function has been associated with anxiety, depression, and other psychiatric conditions (43), with odor information primarily processed in the amygdala, a key stress-response region of the brain (44). Evolutionarily, olfaction aids threat detection, social communication, and survival. Higher anxiety levels have been linked to increased olfactory sensitivity (45), and women with anxiety report greater odor awareness than men (46), consistent with our findings that olfactory transduction-related genes are enriched only in females.

As for the male-specific enrichment in lipoprotein metabolism, both acute and chronic stress lead to elevated levels of lipoprotein in humans. This increase has been explained as a part of the body’s response to provide energy for the fight-or-flight stress-coping mechanism (47). Higher lipoprotein levels have been found to be associated with depression in males (48), while females with higher lipoprotein levels were more likely to have lower scores on depression and anxiety tests (49). These differences may be due to the different lipoprotein metabolism in females and males, which is influenced by estrogen and testosterone-specific signaling pathways (50). The gene enrichment results suggest that genetic factors may contribute to these previously observed sex differences.

Moreover, the male-specific enrichment of the lipoprotein metabolism pathway aligns with our genetic correlation results, which showed a stronger association between waits-hip-ratio adjusted BMI (WHRadjBMI) and anxiety in males. WHRadjBMI reflects central fat distribution and is more closely linked to insulin-related dysregulation and lipoprotein metabolism than BMI (51), thereby providing a plausible mechanism for male-specific anxiety risk. This interpretation is consistent with findings from a large Norwegian population-based cohort, which reported that WHR was associated with anxiety in men but not women (52). Interestingly, BMI showed a stronger genetic correlation with anxiety in females, consistent with epidemiological studies reporting that obesity and anxiety are more strongly linked in women (53,54). A neuroimaging study further showed that higher BMI was associated with significantly stronger stress-related brain responses in females (55), whereas an epidemiological study of young adults found that higher perceived stress was linked to lower BMI in men (56). Overall, these findings suggest that different adiposity pathways underlie sex-specific genetic overlaps between anxiety and body weight, with overall adiposity being more relevant in females and central fat distribution in males.

The genetic correlation between females and males was high but significantly different from one, indicating that the genetic architecture of anxiety is largely shared but not completely identical across sexes. In sex-specific PGS analyses with matched sample size, the only significant difference was that female-derived PGS predicted female anxiety more strongly than male-derived PGS in the population cohort. This finding is consistent with higher SNP heritability observed in females and the expectation that same-sex PGS should align more closely with the target phenotype. In males, female- and male-derived PGS performed similarly, with female-PGS performing slightly better. This may reflect the considerable similarity in genetic effects between sexes and lower SNP-based heritability observed in males. In the QSkin population cohort specifically, anxiety was assessed with a single broad self-report question (‘have you been diagnosed, experienced, or treated for anxiety?’) (35). Such single-item measures have only moderate sensitivity and specificity compared with multi-item scales (57). Underreporting among men (10) may have further contributed to weaker prediction in males relative to other cohorts. In the clinical depression cohort, where anxiety was assessed using DSM-5 criteria (37), and in the GAD-7 symptom-based analyses, PGS showed the expected pattern of stronger same-sex prediction. Although none of these differences reached statistical significance, this aligns with previous studies of sex-specific PGS in depression, which similarly reported no significant differences despite observing the same pattern (58). Taken together, these findings suggest that while subtle sex-specific patterns can be observed, the sex-specific PGS are comparable in predictive ability.

We explored genetic correlations of our sex-specific anxiety GWASs with sex-combined GWAS of other traits, finding that some traits were more strongly correlated in one sex. Bipolar disorders, characterized by alternating manic and depressive episodes, showed a higher genetic correlation with anxiety in type II (depressive episodes are predominant) than type I bipolar disorder (distinguished by manic episodes). While the sex difference in genetic correlation with bipolar disorder did not survive FDR correction (FDR-adjusted P=0.064), it was nominally significant, with males showing stronger correlations across both subtypes. These findings suggest a potential sex-specific genetic relationship between anxiety and bipolar disorder. Given that no consistent sex differences have been observed in bipolar disorder (59), our findings may offer a new perspective on the relationship between anxiety and bipolar disorder through a sex-specific approach. ADHD, despite being more prevalent in males and showing a similar genetic architecture across sexes (60,61), displayed a significantly stronger genetic correlation with female anxiety. This finding is consistent with ADHD and anxiety being more frequently comorbid in females (62). It further implies that in females, ADHD risk may be more likely to manifest through anxiety symptoms, which can result in anxiety diagnoses overshadowing ADHD and lead to under-recognition of ADHD in females (63).

Overall, this study provides evidence of genetic sex differences in anxiety at many aspects, including sex specific variants, prioritized genes, enriched biological processes, genetic architecture, and genetic correlation with other traits. Our findings indicate that sex not only shapes the genetic architecture of anxiety itself but also influences its genetic overlap with comorbid psychiatric conditions. Standard GWAS often assume a shared genetic architecture across sexes, potentially masking sex-specific effects. This was illustrated in the WHRadjBMI analyses, where sex-specific genetic correlations with anxiety were not detectable in the combined GWAS. Given that sex heterogeneity is common in complex traits, ignoring these differences may oversimplify the underlying biology. Future studies conducting sex-specific GWAS on a larger scale are essential for fully capturing the distinct genetic contributions to anxiety in males and females.

Our study is the first to conduct a large-scale sex-stratified GWAS of anxiety, demonstrating significant sex differences in heritability and genetic correlations with other traits. These findings highlight the value of a sex-specific approach for uncovering genetic architecture that may be obscured in combined analyses. However, our results should be considered in light of some limitations. The smaller male sample size reduces statistical power compared to females, and larger samples will be needed to clarify sex differences or commonalities. Additionally, the UK Biobank and All of Us cohorts are not fully representative of the general population, which may limit generalizability. Future sex-specific GWAS in other traits with known heterogeneity could enhance understanding of sex-specific genetic factors and shared genetic architecture across traits.

## Data Availability

All data produced in the present study are available upon reasonable request to the authors

## Acknowledgements

J.H was funded by a QIMR PhD scholarship. B.L.M (APP2017176), K.L.G (APP1173025), S.E.M (APP1172917 and APP2025674), and D.C.W (APP2026567) are supported by Investigator Grants from the National Health and Medical Research Council of Australia (NHMRC). E.M.B are supported by Research Excellence grant (1198304) from NHMRC Centre, and the University of Queensland Health Research Accelerator Program. The QSkin Study is supported by a Clinical Trials and Cohort Grant [APP1185416] from NHMRC. PISA was funded by an NHMRC Dementia Research Team Grant [APP1095227]. This study makes use of data from UK Biobank (Project ID: 25331).

We sincerely thank all UKB, All of Us, AGDS, PISA and QSkin participants for their time and contributions to this study. We also appreciate everyone involved in the study’s conception, implementation, media campaign, and data cleaning.

## Author contributions

J.H performed the analyses, wrote the manuscript and integrated input and feedback from all co-authors. B.L.M and K.L.G designed and supervised the study. Support and input was provided by S.E.M, who along with M.K.L, E.M.B, N.G.M, D.C.W, C.M.O were responsible for the cohort design, genotyping, and data acquisition, processing and quality control. J.T.T provided support with analyses. All authors read and approved the final manuscript.

