## Supplementary Information for "Unravelling Sex Differences in the Genetic Architecture of Anxiety"

Phenotype definition

In this study, we identified participants who met DSM-5 criteria for the following anxiety disorders (1): agoraphobia, social phobias, specific phobias, generalized anxiety disorders, panic disorders and other phobic anxiety disorders.

Phenotype in the UK Biobank

Lifetime anxiety cases in the UKB were defined as individuals that met any of the four following criteria:

i) Individuals who reported having been diagnosed with at least one anxiety disorder by a professional using ICD10 criteria;

ii)  Individuals diagnosed with an anxiety disorder by a professional;

iii) Individuals who met the anxiety criteria of the Composite International Diagnostic Interview (CIDI) short-form questionnaire (2);

iv) Individuals whose total sum score was above or equal to eight in the 7-item general anxiety disorder (GAD-7) scale (3).

We chose eight as the cut-off for the GAD-7 as this has been shown to give the best performance in terms of sensitivity and specificity for classifying anxiety disorders (4). For participants who answered the GAD-7 on multiple occasions, we used the highest sum score as their final score. Among the 15,910 participants with a GAD-7 score ≥ 8, 65.43% (n = 10,411) also met criteria for anxiety based on at least one of the other definitions. The detailed questions and related data fields in the UK Biobank are listed in Table S1.

Participants were classified as controls if they answered no to the following questions *“In your life, have you suffered from a period of mental distress that prevented you from doing your usual activities?”* and *“In your life, did you seek or receive help from a professional (medical doctor, psychologist, social worker, counselor, nurse, clergy, or other helping professional) for mental distress, psychological problems or unusual experiences?”*. In addition, we screened out participants who had been diagnosed with any mental health conditions by a professional or with an ICD10 code, or with a GAD-7 sum score greater than seven.

Phenotype in the All of Us

Lifetime anxiety cases in the AoU dataset were defined as individuals who either had a documented diagnosis in their electronic health record (EHR) of generalized anxiety disorder, panic disorder, or phobic disorder, or who selected “self” in response to the survey question: “Including yourself, who in your family has had anxiety reaction/panic disorder?”

Among females, 17,064 were identified from EHR records and 27,892 from self-report, including 6,996 who met both criteria. Among males, 6,848 were identified from EHR records and 8,043 from self-report, including 1,986 who met both criteria.

Controls were defined as participants who had both EHR and genotyping data available, but did not have any documented EHR diagnosis corresponding to the following conditions:  Major depressive disorder (ID: 440383), Anxiety (ID: 441542), Bipolar disorder (ID: 436665), Schizophrenia (ID: 435783), or Mood disorder (ID: 444100).

GWAS of qualitative GAD-7 in the UK Biobank

We conducted a separate GWAS of the GAD-7 quantitative score in order to evaluate the consistency of this measure with our binary lifetime anxiety phenotype. Among 191,028 participants who completed all seven items, we excluded those with a score of 0 (leaving 106,944) due to the zero-inflated distribution and to focus on individuals with at least minimal anxiety symptoms. Scores were rank-based inverse-normal transformed prior to association testing in REGENIE (v2.2.4) (5).

GWAS was also performed for the other binary definitions of anxiety. Genetic correlation analysis using LD Score Regression (LDSC) (6) demonstrated a high correlation between the quantitative GAD-7 phenotype and the binary lifetime anxiety definition  (rg=0.84 (0.04), P=4.59^-118^).

GWAS of EHR-based anxiety cases in AoU females

In the AoU dataset, more anxiety cases were identified from self-reports than from EHR records. To assess whether including self-reported cases biased the results, we performed a GWAS restricted to female participants with EHR-based diagnoses using REGENIE. We then estimated the genetic correlation between this EHR-only GWAS and our finalized female GWAS in AoU, which included both EHR- and self-reported cases. The correlation was very high (rg=0.95 (0.03), P=8.65^-177^), supporting the validity of including self-reported cases in the final analysis.

Polygenic scores

To test whether our results were able to significantly predict anxiety in an independent sample, we calculated polygenic scores (PGS) using SBayesR (7). SBayesR is a Bayesian technique that estimates the effect of SNPs from multi-normal distributions which may reflect the true distribution of genetic variants. The estimated PGS were standardized using the *scale()* function in R (v4.2.0).

We tested the predictive ability of PGS derived from our GWAS results in the QSkin Sun and Health Study (QSkin). QSkin is a population-based cohort focused on skin cancer and melanoma in Queensland, Australia (8). Participants in QSkin completed online questionnaires, and 2,341 individuals who responded 'yes' to the question 'Have you ever been diagnosed with, experienced or been treated for anxiety?' were classified as cases, and 12,722 individuals with no history of psychiatric disorders were used as controls.

To control for potential familial relationships in the QSkin dataset, we conducted a restricted maximum likelihood (REML) analysis using Genome-wide Complex Trait Analysis (GCTA) (v 1.91.7). We estimated the variance in lifetime anxiety status explained by sex-specific GWAS-derived PGS (PGS_F_ for females and PGS_M_ for males) separately in 8,009 females (N cases = 1,528) and 7,057 males (N cases = 819). We adjusted the model for the first 10 PCs. Similarly, we estimated PGS_C_ from the combined GWAS that included females and males to predict anxiety in all 15,066 individuals, with the model further adjusted for sex as a covariate.

To examine whether our PGS results were influenced by differences in sample sizes, and thus statistical power, across our discovery GWASs, we downsampled the number of cases and controls in the female-specific GWAS to match those in the male-specific GWAS in both UKB and AoU. These downsampled female GWAS results were then meta-analysed and used to generate PRS. To ensure comparable evaluation of PGS predictions, we also downsampled the female prediction cohorts to the same sample size as males (QSkin, AGDS/QSkin, PISA, and AGDS). Downsampling was performed through random selection using the *sample()* function in R(v4.2.0). We then repeated the regression analyses using these downsampled results, following the same procedure as above.

Additionally, we tested the association between PGS_C_, PGS_F_ and PGS_M_ with a continuous measure of current anxiety symptoms, as measured by the GAD-7. The GAD-7 data were available for 4,868 individuals (3,263 females and 1,605 males) in the Prospective Imaging Study of Ageing: Genes, Brain and Behaviour (PISA) (9). PISA is an Australian-based cohort of middle-aged and older participants investigating risk factors and biomarkers for dementia. Participants completed an online survey which included the GAD-7. Associations of PGS and GAD-7 were tested using the REML approach in GCTA, similar to the analysis of anxiety in QSkin.

Finally, we tested the PGS predictions of both lifetime anxiety and current anxiety (GAD-7) in a secondary sample, the Australian Genetics of Depression Study (AGDS). AGDS is an Australia-based cohort, including approximately 17,000 genotyped participants (10) that report a history of depression. Participants in AGDS completed online questionnaires, and those who reported a diagnosis of generalized anxiety disorders were classified as lifetime anxiety cases, and the individuals reported no history of psychiatric disorders from QSkin were used as the controls. There are 14,759 females (cases=8,278) and 9,140 males (cases=2,902) for prediction of lifetime anxiety. GAD-7 data were available for 7,282 individuals (5,556 females and 1,681 males) in AGDS. While larger in size, it is important to note that in AGDS all the anxiety cases have comorbid depression. Therefore, this sensitivity analysis may distinguish prediction between population-based, non-comorbid cohorts and clinical, comorbid-depression cohorts. All models were adjusted for the same covariates as in the approach above and GCTA was used for analyses.

Supplementary Figures

Figure S1: Manhattan plot of sex-combined and sex-specific GWAS in the UK Biobank

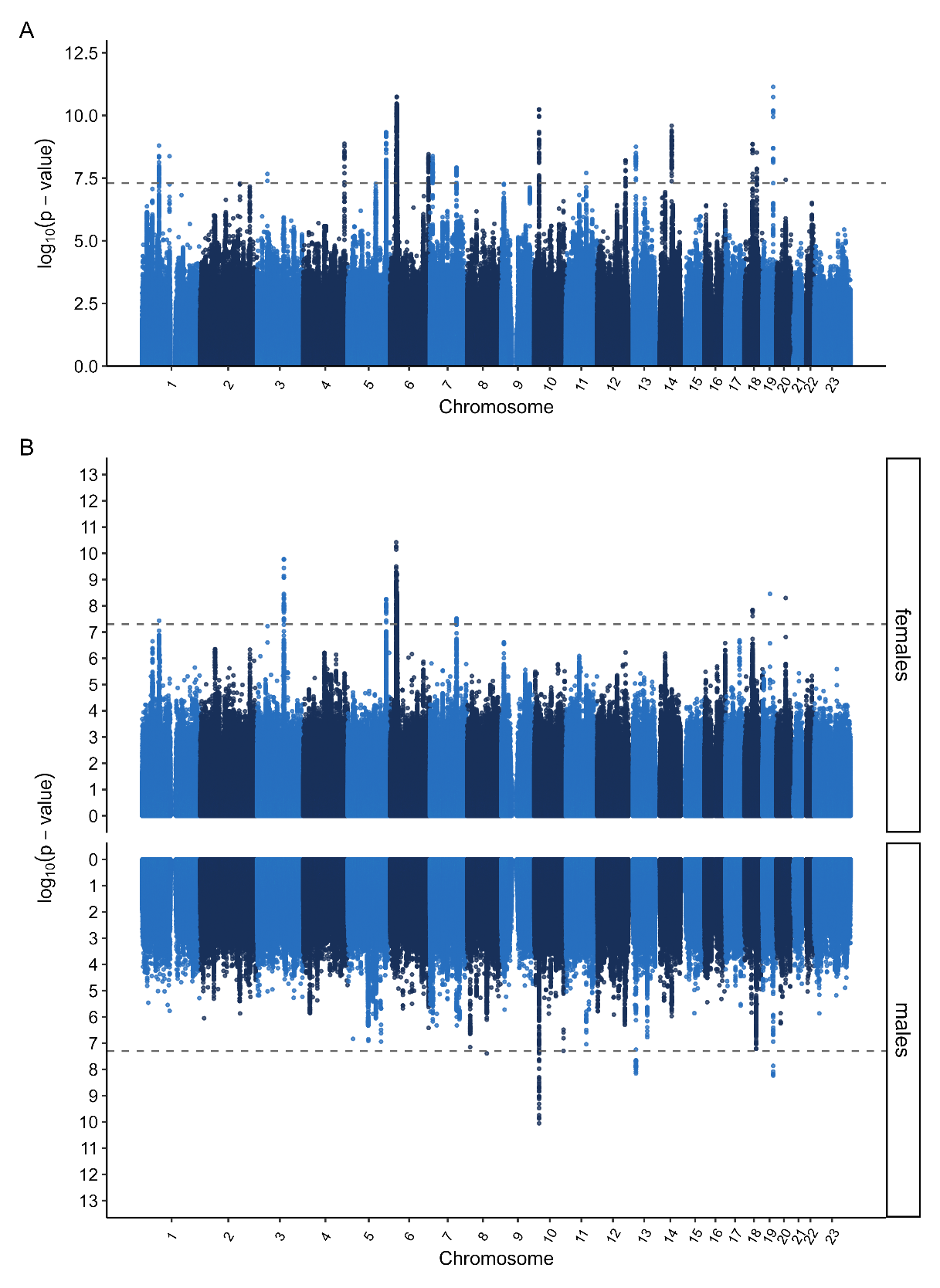

**Figure S2:  Manhattan plot of sex-combined and sex-specific GWAS in the All of Us**

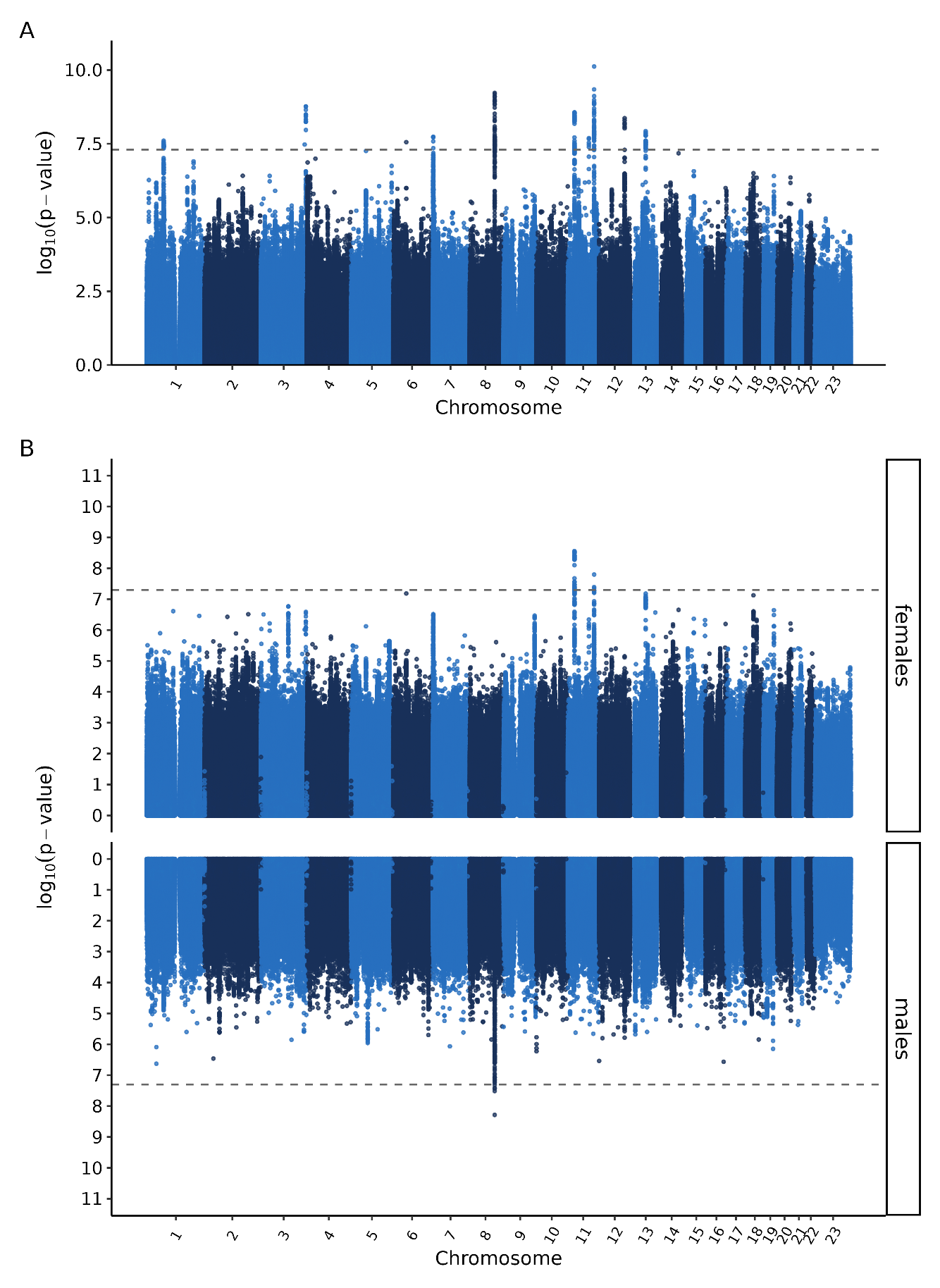

Figure S3: Sex-specific polygenic risk scores (PRS) in the clinical sample.

**3A**: PGS prediction for lifetime anxiety in AGDS/Qskin. **3B**: PGS prediction of GAD-7 in AGDS.

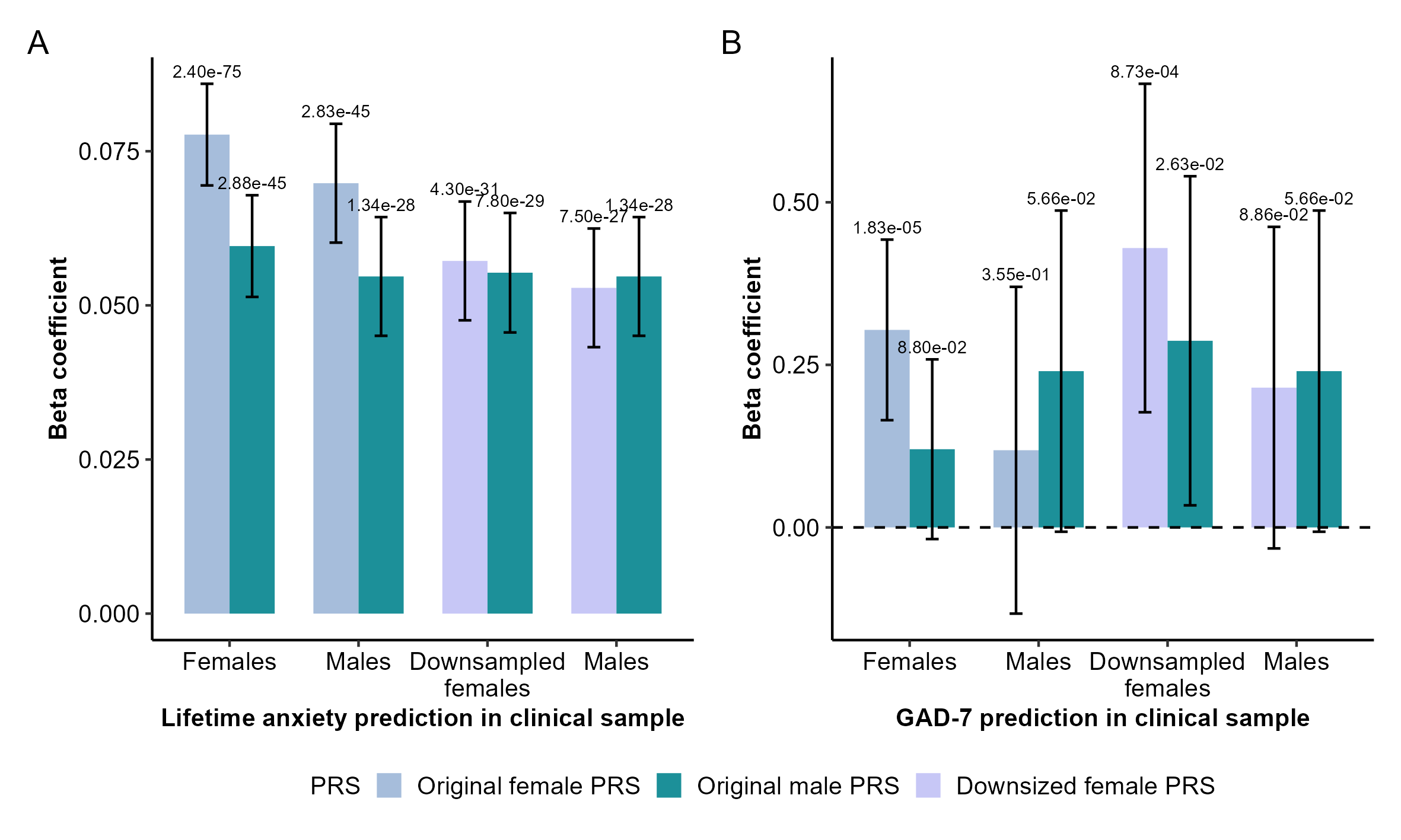
